# City-wide wastewater genomic surveillance through the successive emergence of SARS-CoV-2 Alpha and Delta variants

**DOI:** 10.1101/2022.02.16.22269810

**Authors:** F.S. Brunner, M.R. Brown, I. Bassano, H. Denise, M.S. Khalifa, M. Wade, J.L. Kevill, D.L. Jones, K. Farkas, A.R. Jeffries, The COVID-19 Genomics UK (COG-UK) Consortium, E. Cairns, C. Wierzbicki, S. Paterson

**Author notes:** https://www.cogconsortium.uk, Full list of consortium names and affiliations are in the appendix (The COVID-19 Genomics UK (COG-UK) Consortium). These authors contributed equally to the project (F.S. Brunner, M.R. Brown).

## Abstract

Genomic surveillance of SARS-CoV-2 has been essential to provide an evidence base for public health decisions throughout the SARS-CoV-2 pandemic. Sequencing data from clinical cases has provided data crucial to understanding disease transmission and the detection, surveillance, and containment of outbreaks of novel variants, which continue to pose fresh challenges. However, genomic wastewater surveillance can provide important complementary information by providing estimates of variant frequencies which do not suffer from sampling bias, and capturing all variants circulating in a population. Here we show that genomic SARS-CoV-2 wastewater surveillance can detect fine-scale differences within urban centres, specifically within the city of Liverpool, UK, during the emergence of Alpha and Delta variants between November 2020 and June 2021. Overall, the correspondence between wastewater and clinical variant frequencies demonstrates the reliability of wastewater surveillance. Yet, discrepancies between the two approaches when the Alpha variant was first detected emphasises that wastewater monitoring can also capture missing information resulting from asymptomatic cases or communities less engaged with testing programmes, as found by a simultaneous surge testing effort across the city.

## Introduction

Genomic surveillance has been a significant feature in the public health response to the SARS-CoV-2 pandemic (Wu et al. 2020; Zhu et al. 2020) because of its ability to detect the emergence of and track new variants of concern (VOC) (Robishaw et al. 2021; Vöhringer et al. 2021). Important examples include the B.1.1.7 (Rambaut et al. 2020), B.1.351 (Tegally et al. 2020), P.1 (Faria et al. 2021), B.1617.2 (Cherian et al. 2021) and, most recently, the B.1.1.529 lineage (Qin et al. 2021), named VOC Alpha, Beta, Gamma, Delta, and Omicron, respectively. These VOC have demonstrated one or more attributes out of increased transmissibility, more severe disease, a reduction in antibody neutralisation, reduced therapeutic response or reduced vaccine effectiveness (Davies et al. 2021; Robishaw et al. 2021). Thus, while vaccination currently provides substantial protection against all known VOC, continued genomic surveillance is essential to mitigate and contain the threat they pose to public health. It informs the implementation and assessment of non-pharmaceutical interventions (e.g. social distancing, lockdowns and regional, national and international restrictions) and targeted surge testing, and serves as an early warning system for the emergence and spread of novel variants (Mishra et al. 2021; Tegally et al. 2020). Genomic surveillance of SARS-CoV-2 has primarily been driven by whole genome sequencing of clinical isolates, typically using residual RNA from diagnostic RT-qPCR tests. One million SARS-CoV-2 genomes were sequenced worldwide by April 2021, rising to over seven million by January 2022 on the GISAID database (Elbe and Buckland-Merrett 2017). This has provided unprecedented insight into the joint evolution and epidemiology of the SARS-CoV-2 pandemic (Ward et al. 2021; Harvey et al. 2021). Nevertheless, the cost of clinical sequencing to generate these data has been, and continues to be, substantial (10 - 35 GBP per sample for consumables (Tyson et al. 2020), plus similar costs for staff, logistics and data infrastructure). It may be unsustainable at the levels required to adequately inform public health authorities as SARS-CoV-2 becomes endemic and threatens public health for the foreseeable future, even in developed nations.

Wastewater-based surveillance is a complementary, cost-effective approach to clinical sequencing, which has gained significant attention throughout the COVID-19 pandemic (Jahn et al. 2021; Hillary et al. 2021; Peccia et al. 2020; Smyth et al. 2021; Mishra et al. 2021). Given that SARS-CoV-2 is shed in faeces by more than 50% of infected people (Foladori et al. 2020), it can be recovered from wastewater, its RNA extracted, and its presence and quantity in a wastewater catchment determined using RT-qPCR (Farkas et al. 2020), with trends generally tracking the rise and fall of corresponding clinical cases (Peccia et al. 2020; Hillary et al. 2021; Wade et al. 2020). This can be achieved for entire populations by sampling at the inlet of wastewater treatment plants, or at much finer spatial scales, such as across cities, by sampling within the sewer network.

More recently, the recovery of SARS-CoV-2 genomes from wastewater has opened up the possibility of detecting and tracking circulating SARS-CoV-2 variants (Jahn et al. 2021; Hillary et al. 2021; Peccia et al. 2020; Smyth et al. 2021; Brown et al. 2021). Such an approach is particularly attractive for population-level insights during periods of high prevalence, especially if capacity constraints reduce the proportion of sequenced positive RT-qPCR tests. Furthermore, it can detect asymptomatic cases and is proposed to capture communities under-represented by clinical testing, particularly in urban centres (Polo et al. 2020; Green et al. 2021). Nevertheless, moving from detection and quantification of SARS-CoV-2 in wastewater by RT-qPCR to characterisation by genome sequencing is challenging.

The low abundance of SARS-CoV-2 means enrichment through RNA concentration methods is necessary, simultaneously enriching PCR inhibitors and contaminating bacterial, viral, and human nucleic acids (Peccia et al. 2020). SARS-CoV-2 genomes in wastewater are also highly degraded and fragmented. In combination, this can result in poor and inconsistent amplification of target amplicons and thus patchy genome coverage. Even if amplification and sequencing are successful, data interpretation can be difficult. Wastewater harbours a mixed SARS-CoV-2 population. Therefore, sequences are derived from a pool of fragments, removing much of the phase information between polymorphic sites on the genome used to assign phylogeny and subsequently lineage. However, by reference against clinically-derived genomes of known SARS-CoV-2 lineages, wastewater data has the potential to detect and quantify polymorphisms characteristic of defined lineages and of VOC in particular (Jahn et al. 2021; Fontenele et al. 2021).

This study demonstrates the utility of wastewater-based genomic surveillance of SARS-CoV-2 using longitudinal data collected from multiple locations in a single city – Liverpool, UK – between November 2020 and June 2021. During this time, Liverpool was the subject of a pilot study evaluating lateral flow tests for rapid asymptomatic testing (García-Fiñana et al. 2021). This pilot noted the link between social inequalities and testing uptake, with social deprivation and digital exclusion as major factors limiting uptake (Green et al. 2021). Wastewater-based epidemiology can provide valuable insight into some of the communities or areas of Liverpool that may be less accessible to conventional testing. This period in the UK also saw the emergence and establishment of Alpha (B.1.1.7) and subsequently Delta (B.1.617.2) SARS-CoV-2 variants (Vöhringer et al. 2021). We show that wastewater genomic surveillance can reliably detect the emergence of these variants and their subsequent rise across a city.

## Methods

### Sample Collection, Concentration and RNA Extraction

Wastewater grab samples (1 L per sample) were collected from eight locations across the Liverpool sewer network and from Liverpool’s main wastewater treatment plant at Sandon Docks (WWTP) between the 2^nd^ of November 2020 until the 21^st^ of June 2021, as part of the ongoing Environmental Monitoring for Health Protection programme (part of NHS Test & Trace, now the UK Health Security Agency) in England (Fig. 1, Table S1). In addition, concurrent samples from four WWTPs in the southeast of England were collected between 2^nd^ of September 2020 and 17^th^ of January 2021 as a control group. Samples were transported and subsequently stored at 4 - 6°C until analysis, minimising RNA degradation. Within 24 h of collection, all samples were centrifuged (10,000 x g, 4 °C, 10 min) in sterile polypropylene tubes to remove suspended solids. The supernatant (50 ml) was transferred to 250 ml polycarbonate PPCO bottles containing 19-20 g of ammonium sulfate (Sigma-Aldrich, Cat. No. A4915). After the ammonium sulfate had dissolved, the samples were incubated at 4 °C for 1 h before further centrifugation (10,000xg, 4 °C, 30 min) and supernatant removal. The pellet was resuspended in 200-500 _μ_l of PBS. Concentrates were stored at 4 °C until nucleic acid extraction. Nucleic acids were extracted from concentrates using NucliSens lysis buffer (BioMerieux, Marcy-lÉtoile, France, Cat No. 280134 or 200292), NucliSens extraction reagent kit (BioMerieux, Cat. No. 200293) either manually (Farkas et al. 2021) or using the King-fisher 96 Flex system (Thermo Scientific, Waltham, MA, USA) according to the manufacturer instructions (Kevill et al. 2022), generating RNA extracts of 50 - 100 µL in volume. Extracts were stored at -80°C until further processing.

**Figure 1.**
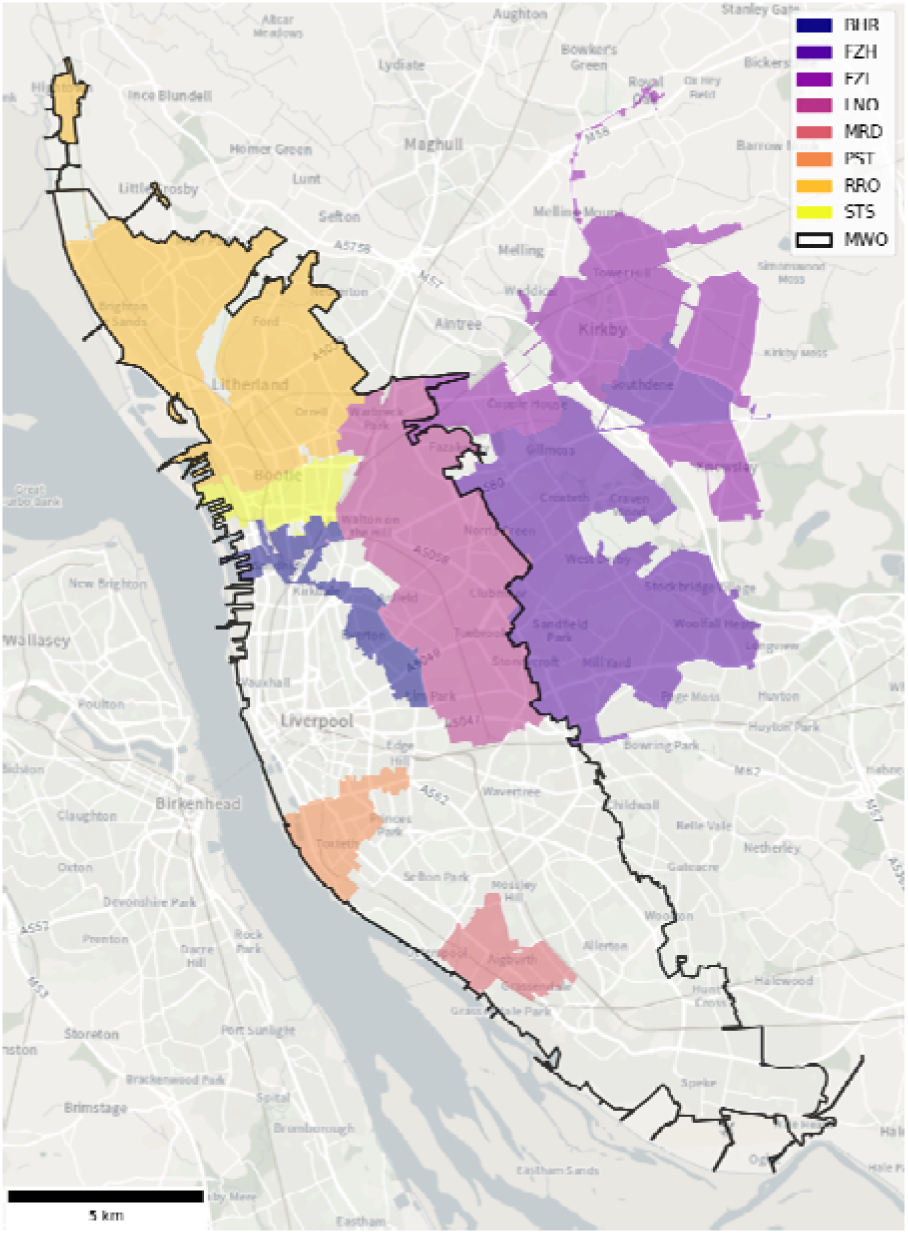
Wastewater catchments of the 8 sewer network sampling locations (coloured) and the WWTP (MWO, black outline) across Liverpool. BHR= Bank Hall Relief, FZH= Fazakerley High, FZL= Fazakerley Low, LNO= Liverpool North, MRD= Mersey Road, PST= Park Street, RRO= Rimrose, STS= Strand SSO, MWO= Sandon Dock Main Works. See Table S1 for further catchment details.

### SARS-CoV-2 RNA Amplicon Sequencing

Wastewater RNA extracts were purified and sequenced with a standardised EasySeq™ RC-PCR SARS CoV-2 (Nimagen) V1.0 protocol (Jeffries et al. 2021). In short, samples were cleaned with Mag-Bind® TotalPure NGS beads (Omega Bio-Tek) and then reverse transcribed using LunaScript® RT SuperMix Kit (New England Biolabs) and the EasySeq™ RC-PCR SARS CoV-2 (novel coronavirus) Whole Genome Sequencing kit v3.0 (NimaGen). Amplicons were pooled and libraries cleaned with Mag-Bind® Total Pure NGS beads (Omega Bio-Tek) before sequencing on an Illumina NovaSeq™ 6000 platform generating 2×150bp paired end reads.

### Bioinformatics Analysis

We processed raw reads using the *ncov2019-artic-nf v3* pipeline (https://github.com/connor-lab/ncov2019-artic-nf) using default parameters. Briefly, reads were aligned to the SARS-CoV-2 reference genome (MN908947.3, (Wu et al. 2020)) using the Burrow-Wheeler Aligner (bwa) (Li and Durbin 2009). We then identified Single Nucleotide Polymorphisms (SNPs) and insertions/deletions (Indels) from BAM files using samtools (v1.13, (Danecek et al. 2021)) and VarScan (v2.4.4, *P* < 0.05, all other settings default, (Koboldt et al. 2012)) on 100,000 sequencing reads with an alignment score > 10. Next, we filtered the identified SNPs and Indels against signature mutations of known VOC and variants under investigation (VUI), as defined by Public Health England (PHE) at the time of writing (https://github.com/phe-genomics/variant_definitions, Table S2, Fig. 2). We further used custom scripts to extract summary statistics and sequence quality indicators, such as genome coverage, mapped reads and read depth (Table S3).

**Figure 2.**
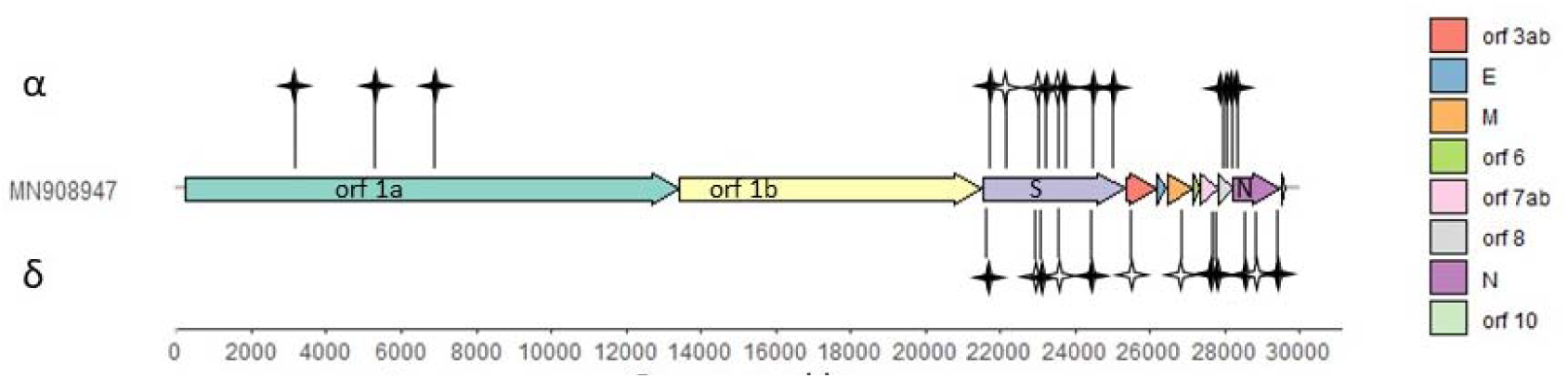
SARS-CoV-2 genome structure and signature mutations of VOC Alpha and Delta. Black stars show unique mutations for the Alpha or Delta variant. White stars show mutations shared with at least one other VOC or VUI and therefore not used for mean frequency estimates. Details of all mutations can be found in Table S2. Genome structure adopted from (Wu et al. 2020).

To aid VOC and VUI identification at low frequencies from wastewater samples, we adopted a recently described amplicon-level co-occurrence approach (Jahn et al. 2021). Briefly, co-occurring mutations were called from BAM files using CoOccurrence adJusted Analysis and Calling (COJAC) (Jahn et al. 2021), facilitating the identification of signature mutations co-occurring on the same sequencing read, that is, a read or paired read coming from the same amplicon, thus one SARS-CoV-2 virion. This greatly improves confidence in variant detection, especially at low frequencies, since co-occurring mutations are less likely to arise through sequencing error than individual SNPs (Jahn et al. 2021). We extracted co-occurrence signature mutations of the B.1.1.7 (VOC-20DEC-01, alpha) and B.1.617.2 (VOC-21APR-02, delta) lineages. Since a proportion of signature mutations are shared amongst VOC/VUI, not all variants have a unique set of co-occurring mutations. B.1.1.7 has one unique pair of co-occurring mutations on amplicon 147, while B.1.617.2 only has one non-unique pair of mutations on amplicon 121, i.e., it is shared with other variants.

### Statistical Analyses

We used RStudio v. 1.4.1717 (RStudio Team 2021) and R version 4.1.1 (R Core Team 2021) for all statistical analyses and ggplot2 (Wickham 2016) for visualisations. Prior to analysis, all unique signature mutations (SNPs/Indels) of a given variant (Fig. 2) were identified in each sample, mutations with a read depth <10 removed and frequencies of 1.0 and 0 rescaled to 0.99 and 0.01 for beta regression compatibility, respectively. We modelled the relationship of the mean frequency of each variant’s signature SNPs/indels with location (i.e. differing network sites) and time during respective variant emergences with beta linear regression (betareg, “betareg” v.3.1.4, (Cribari-Neto and Zeileis 2010)), given allele frequencies are in the standard unit interval [0, 1]. To do so, we set the mean frequency of the unique signature SNPs/indels of a given variant as the dependent variable, and wastewater site, date and their interaction as predictor variables. All models were fit by maximum likelihood using the logit link function, *logit(p)* with *p* the probability of observing the (variant) data and *logit* being the inverse of the standard logistic function, and included site as an additional regressor for the precision parameter when it improved the model fit (see Table S4 for final model structures), as indicated by Akaike Information Criterion (AIC) and likelihood ratio tests (Cribari-Neto and Zeileis 2010). To account for missing data in SNP/indel frequencies, a weighting factor was applied using the number of used signature SNPs/indels (weights = *n*). We assessed model validity by visual checks of homoscedasticity of the standardised weighted residuals and linearity of the model fit (Fig. S2). We then extracted likelihood ratio tests of estimated marginal means for each predictor variable (joint_tests, “emmeans” (Lenth 2021), Table S4).

We also compared the frequency of detected VOC/VUI signature SNPs/indels in wastewater samples to the frequency of VOC/VUI identified in clinical cases by the COVID-19 Genomics (COG-UK) Consortium between the 2^nd^ of November 2020 and the 21^st^ of June 2021 across Liverpool. We extracted counts of genomically confirmed cases for all circulating lineages from the CLIMB platform (Nicholls et al. 2021) on 26^th^ of October 2021 and then filtered and grouped by the outer postcodes covered by the catchment areas of the WWTP and the eight sewer network sites. For the Delta variant, confirmed clinical cases of the B.617.2 lineage and its subvariant AY.4 were combined. Where outer postcodes spanned multiple wastewater catchments, we included clinical cases in counts for all those sites, divided by the number of overlapping wastewater catchments. Additionally, we obtained total daily infection numbers for the upper tier local authority of Liverpool from UK Government statistics (https://coronavirus.data.gov.uk, Fig. S5).

## Results

Across all catchments, we observed a significant increase in the mean frequency of Alpha (B.1.1.7) signature SNPs/indels between the 2^nd^ November 2020 and the 28^th^ February 2021 (F_1_ = 13667, *P* < 0.001, Fig. 3, Table S4). This closely corresponds with the observed rise in Alpha clinical cases across Liverpool for the same period (Fig. 4) and wastewater data from four WWTPs in the southeast of England (F_1_ = 13829, *P* < 0.001, Table S4, Fig. S3). For most sites, the rise of Alpha began in mid to late December, with peak frequencies observed in late January and early February (Figs. 3 and 4). As defined by co-occurrence analysis, the earliest wastewater Alpha detection preceded clinical detections in five of the nine sites by up to 55 days (Fig. 4). The contrary was observed in the remaining four sites, with clinical samples picking up Alpha up to 26 days earlier (Fig. 4).

**Figure 3.**
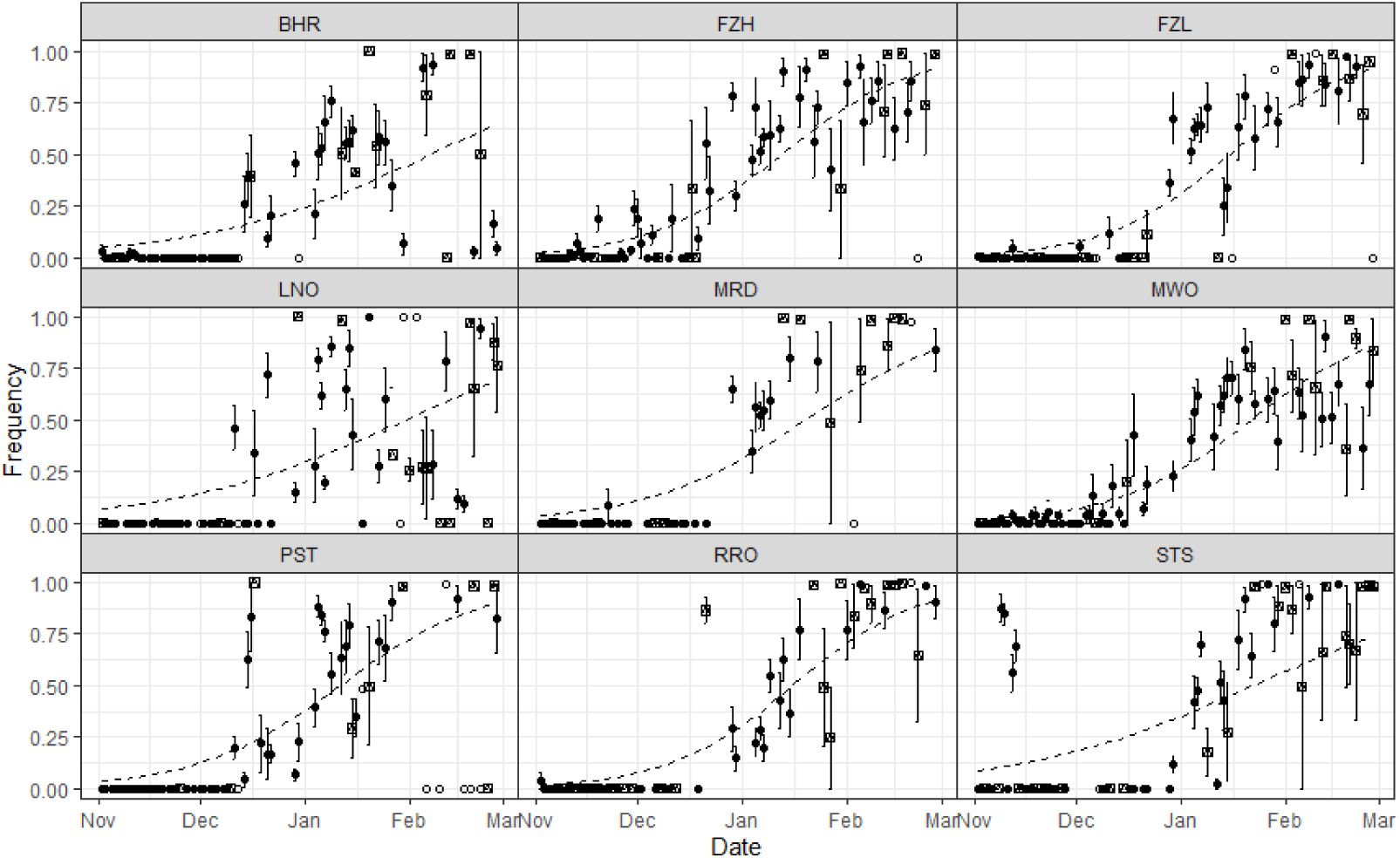
Mean frequency of B.1.1.7 (Alpha) signature SNPs/Indels in each wastewater catchment, 2^nd^ November 2020 to 28^th^ February 2021. Points and error bars show means and standard errors across all unique Alpha mutations with sufficient sequencing coverage in each sample. Dashed lines show the best fit beta regression line for this period (Table S4). Point shape indicates the number of unique alpha mutations used in the mean calculation for a given sample: empty circles: 1 mutation, crossed square: 2 to 5 mutations, filled circles: >5 mutations.

**Figure 4.**
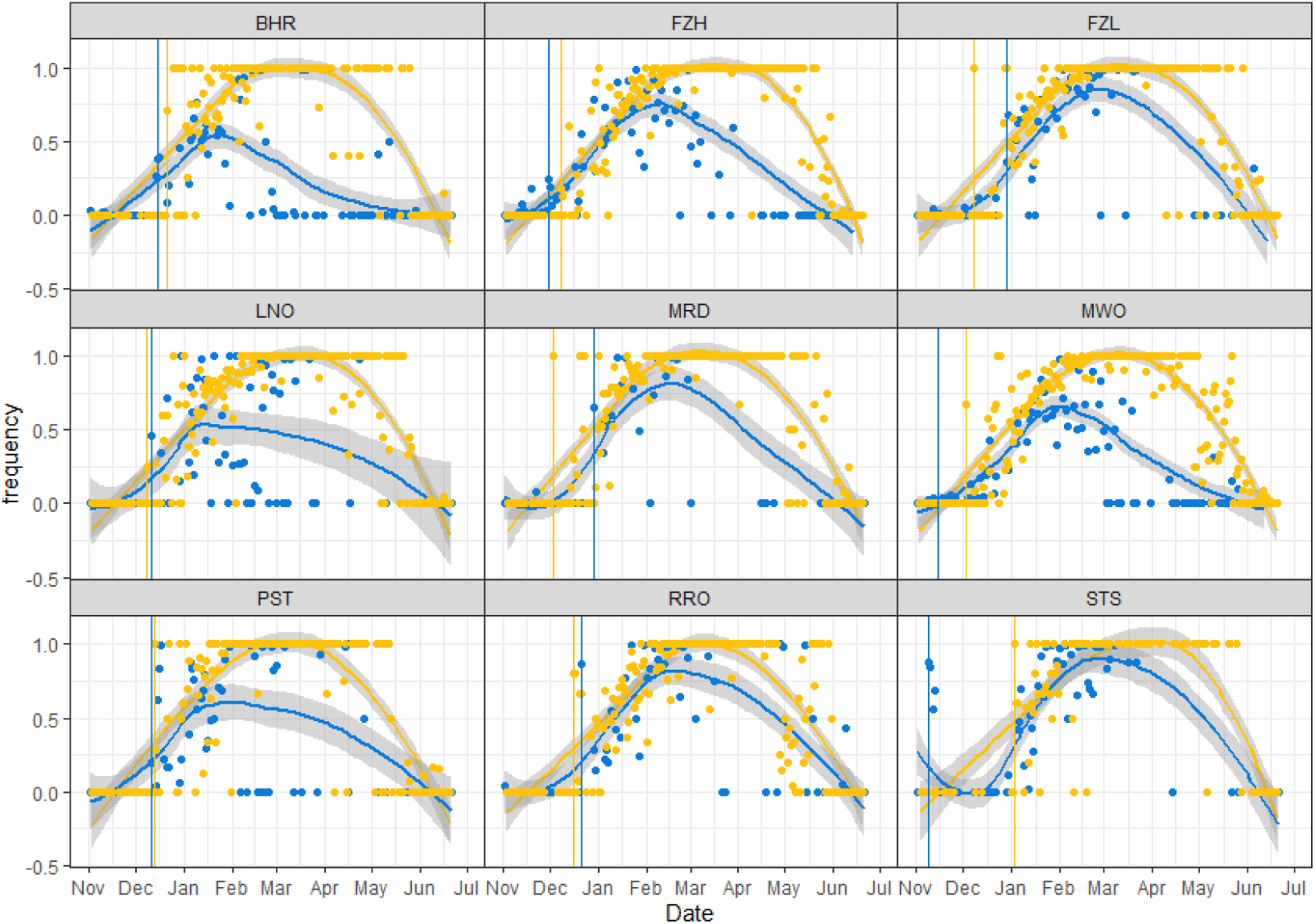
Mean frequency of B.1.1.7 (Alpha) signature SNPs/Indels detected in wastewater versus clinical samples in each catchment, 2^nd^ November 2020 to 26^th^ June 2021. Points show the mean frequency of unique Alpha mutations for a given wastewater sample (blue) and the frequency of Alpha clinical cases from a given date (yellow). Coloured lines show the respective local polynomial regression fit including shaded 95% confidence intervals. Vertical lines indicate the first confirmed clinical case of Alpha (yellow) and the first wastewater detection of co-occurring Alpha mutations on amplicon 147 (blue).

We detected local differences between wastewater catchments (date: site interaction, F_8_ = 32.8, *P* < 0.001, Table S4). This was most notable in Strand SSO (STS), where we observed a high frequency of Alpha signature SNPs from four samples in early November (Fig. 3), though this signal diminished before further detections in early January. Similarly, we observed Alpha signature SNPs at a low to moderate frequency at Fazakerley High (FZH) as early as mid-November, while they were barely detected until late December in Mersey Road (MRD, Fig. 3). This suggests Alpha spread through parts of the north of the city earlier than through the south (Fig. 1), a finding corroborated by co-occurrence analysis but not clinical data (Fig. 4).

From 15^th^ March to 26^th^ June 2021, we observed a significant increase in the frequency of Delta signature SNPs in all wastewater catchments (F_1_ = 964.1, *P* < 0.001, Table S3, Fig. 5), with variation in temporal trends across the city (date: site interaction, F_8_ = 32.4, *P* < 0.001, Table S4, Fig. 5). This coincides with a rise in clinical cases of the same variant (B.1.617.2 and AY.4, Fig. S4) and Alpha’s decline (Figs. 3 and 4). It is noteworthy that the observed transition from Alpha to Delta in wastewater was abrupt (Figs. 4, 5 and S4). From April to early June, infection numbers were low across Liverpool (Fig. S5), and wastewater SARS-CoV-2 concentrations were consequently low (J. Kevill & D. Jones, unpublished data). This is reflected in the observed reduction in mapped reads and genome coverage for this period (Fig. S1). Indeed, the detection of Alpha and Delta signature SNPs was more sporadic during this period (Figs. 4, 5 and S4). Co-occurrence analysis was not informative for the Delta variant since the only pair of signature mutations co-occurring on the same amplicon of our panel is not unique to the Delta variant but also part of the B.1.629 and AY.3 variants, among others. This explains why we detected a co-occurrence signal as early as November for all sites (Fig. S4).

**Figure 5.**
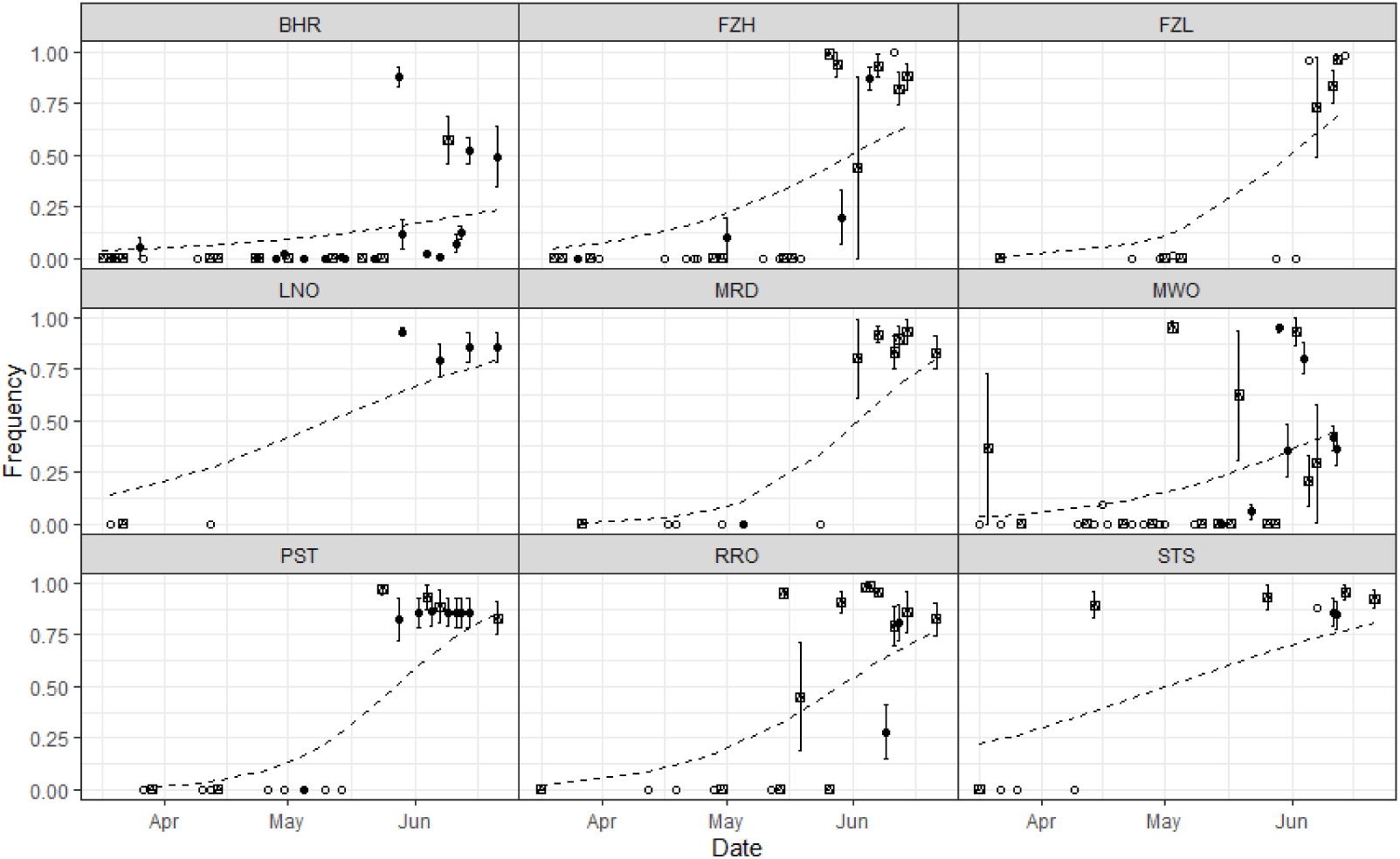
Mean frequency of B.1.617.2 (Delta) signature SNPs/Indels in each wastewater catchment, 15^th^ March 2021 to 26^th^ June 2021. Points and error bars show means and standard errors across all unique Alpha mutations with sufficient sequencing coverage in each sample. Dashed lines show the best fit beta regression line for this period (Table S4). Point shape indicates the number of unique alpha mutations used in the mean calculation for a given sample: empty circles: 1 mutation, crossed square: 2 to 5 mutations, filled circles: >5 mutations.

Finally, we also observed signature SNPs of other VOC and VUI in wastewater samples across Liverpool (Fig. S6), however detections were sporadic and at low predicted frequencies, which corresponds to clinical cases. Although there were notable exceptions, including the B.1.351 (Beta), P.1 (Gamma), P.3 (Theta) and B.1.1.318 lineages (Fig. S6).

## Discussion

Genomic surveillance of wastewater has already shown great promise throughout the unfolding SARS-CoV-2 pandemic (Polo et al. 2020; Farkas et al. 2020), including the detection of VOC (Fontenele et al. 2021), in some cases prior to clinical detection (Jahn et al. 2021). Here, we have demonstrated that wastewater monitoring can also reveal fine-scale, local differences in the spread of VOC across urban centres. Spatiotemporal differences in variant frequencies across Liverpool were recoverable throughout the rise of Alpha (B.1.1.7) in early winter 2020 and, despite lower quality data, the rise of Delta (B.1.617.2/AY.4) in spring 2021. This clear reflection of the rise of Alpha and Delta, respectively, in clinical and wastewater genomic data demonstrates the reliability of this approach.

As seen for the Alpha variant here and by Jahn et al. (2021), genomic surveillance of wastewater can detect VOC earlier than clinical testing. In both instances, co-occurrence analysis improved confidence in (early) low-frequency variant detection by identifying multiple linked mutations from the same virion instead of solely relying on single signature mutations. This requires the co-occurrence of mutations unique to a given variant on amplicons of the used sequencing scheme. When no unique mutation set is available, as is the case for the Delta variant and the NimaGen SARS-COV-2 whole-genome sequencing kit used here, reliable variant detection via co-occurrence analysis is not possible. The software developers have acknowledged this limitation, and the design of primers to create appropriate co-occurrence amplicons for relevant sequencing schemes is suggested as a workaround (Jahn et al. 2021). It is important to note that even in cases where co-occurrence analysis is applicable, our fine-scale local data highlighted that wastewater monitoring sometimes detects new variants earlier than clinical testing, but not always. The reasons for this are yet unclear. It is likely that the inherent variability of wastewater detections, due to variations in viral shedding rates and dilution from rainfall, etc (Polo et al. 2020)), and the increasing stochasticity of clinical detection with decreasing population size play a role. Certainly the relationships between population size, wastewater flow variation and SARS-CoV-2 variant detection warrant further investigation.

This mixed pattern of wastewater Alpha detections preceding confirmed clinical cases in some parts of Liverpool, yet vice versa in others, highlights the complementarity of the approaches. Wastewater monitoring has the notable advantages of being more cost-effective per unit of population and is less biased by testing frequencies in different communities (Polo et al. 2020), while sequencing of clinical samples provides greater specificity and the opportunity for contact tracing. Indeed, our finding that Alpha was detected in wastewater in North Liverpool much earlier than clinical cases had indicated, corresponds well with findings from a large-scale asymptomatic testing campaign, which found that testing uptake was lower in North Liverpool, yet the rate of positive tests higher (Green et al. 2021). Clearly, a combination of genomic surveillance of clinical cases and wastewater is most likely to detect new variants as early as possible and provides the most precise picture of unfolding variant dynamics to inform public health measures (Mishra et al. 2021).

Intriguingly, when comparing peak Alpha and Delta frequencies in corresponding clinical and wastewater data, we find that each variant, in turn, reaches complete dominance, with estimated frequencies of 100% and ∼75% in clinical and wastewater samples, respectively. This suggests improved statistical methods may be required to estimate lineage frequencies from pooled sequencing data, as produced from wastewater samples. Here we have relied on a relatively crude estimation by taking the mean of signature SNP/Indel frequencies. However, genetic variation may mean that a SNP/Indel may not be present on all branches within a lineage, whereas a single fully phased viral genome from a clinical sample would be reliably assigned to a lineage. Following methods for analysing metagenomic amplicon data (Quince et al. 2011), the development of statistical methods to infer lineage proportions from multiple amplicons, while controlling for sequencing error may be productive.

The limited quality of sequences obtained from wastewater during April and May 2020 also highlights the current limits of variant detection via wastewater sequencing when case numbers, and hence SARS-CoV-2 concentrations, are low. While the rise of Delta was evident in our results (Fig. 5), the transition from Alpha to Delta, compared to the gradual emergence of Alpha, was less visible and more abrupt. The use of a relatively short amplicon scheme, as used here, may help to boost sensitivity given the likely degradation of SARS-CoV-2 RNA in wastewater; and from initial work gives better genome coverage than the longer ARTIC v3 amplicons more widely used for sequencing SARS-CoV-2 (data not shown). As previously noted, however, the use of short amplicons limits the use of co-occurrence analysis for variant detection. It is also anticipated that the increased adoption of wastewater-based epidemiology will drive innovation in wastewater sampling, concentration and RNA extraction, improving viral qPCR and sequencing sensitivity (Polo et al. 2020; Hillary et al. 2021; Kevill et al. 2022).

In conclusion, we show that wastewater genomic sequencing can detect the emergence and rise of new SARS-CoV-2 variants. Results correspond well with those obtained through genomic sequencing of clinical samples. In some cases, variants are observed prior to clinical detections, which may be particularly useful in areas or communities with low testing uptake.

## Data Availability

Wastewater sequencing data will be deposited and publicly available on the European Nucleotide Archive by 1st July 2022. The clinical case data used in this study is visualised at https://www.cogconsortium.uk/tools-analysis/public-data-analysis-2/. A filtered, privacy conserving version of the lineage-LTLA-week dataset is publicly available online (https://covid19.sanger.ac.uk/downloads) and gives access to almost all used data, despite a small number of cells having been suppressed to conserve patient privacy.

## Acknowledgements

We thank R. Crompton, B. Jones, G. Airey, T. Foster, N. Kadu, C. Nelson and A. Lucaci for help with processing samples and sequencing and United Utilities for providing wastewater catchment mapping data. Funding was provided by NERC (NE/V003860/1) and DHSC UK (2020_097). COG-UK is supported by funding from the Medical Research Council (MRC) part of UK Research & Innovation (UKRI), the National Institute of Health Research (NIHR) [grant code:MC_PC_19027], and Genome Research Limited, operating as the Wellcome Sanger Institute.

## Data accessibility statement

Wastewater sequencing data will be deposited and publicly available on the European Nucleotide Archive by 1^st^ July 2022. The clinical case data used in this study is visualised at https://www.cogconsortium.uk/tools-analysis/public-data-analysis-2/. A filtered, privacy conserving version of the lineage-LTLA-week dataset is publicly available online (https://covid19.sanger.ac.uk/downloads) and gives access to almost all used data, despite a small number of cells having been suppressed to conserve patient privacy.

## Ethics Statement

Use of surplus nucleic acid derived from routine diagnostics and associated patient data was approved through the COG-UK consortium by the Public Health England Research Ethics and Governance Group (R&D NR0195).

